# Scalable frameworks for local evaluation of AI-enabled imaging devices using PACS-integrated large language models

**DOI:** 10.64898/2026.11.25.26368956

**Authors:** Ashwin Amurthur, Samuel A Justice, John K Chin, Sarah F Mercaldo, Ryan J Morley, Alexander S Schultz, Vasco Silva, Tae Jin Yun, Jin Wook Choi, Christopher P Bridge, Thomas Schultz, James M Hillis

## Abstract

**Introduction:** Artificial intelligence (AI)-enabled imaging device performance may vary across institutions due to differences in disease prevalence, patient demographics, and acquisition protocols. Routine local evaluation, including pre-deployment baseline testing and post-deployment monitoring, remains limited by manual data curation. A large language model (LLM)-as-a-judge approach using existing radiology reports provides a method of performing local evaluation.

**Methods:** We established PACS-integrated LLMs in HIPAA-compliant environments. We then developed two scalable frameworks to automate key processes of local evaluation: selection of a candidate series for device inference and generation of report-derived labels. We applied these frameworks to 4,000 consecutive non-contrast head CT examinations from two hospitals to evaluate agreement between radiology reports and Purple AI Hyper Insight-ICH, a commercial intracranial hemorrhage detection device.

**Results:** For series selection, we used a three-tiered, ‘traffic-light’ approach across 31,422 available series to select a single series for device inference in 3,976 examinations. For radiology report field extraction, we refined an LLM prompt with three steps: initial binary classification, characterization of discordance between the binary classification prompt output and device output, and a comprehensive prompt (featuring 28 structured fields that were informed by the preceding stages). The positive percent agreement (PPA) depended on the definition of positive cases based on the comprehensive prompt fields: it was 87.7% (79.2 to 95.0%) using similar criteria to many published studies (acute or acute-on-chronic, non-minimal, non-equivocal hemorrhage), yet was lower when including chronic, minimal or equivocal ICH. The negative percent agreement (NPA) was 94.7% (94.0 to 95.4%). Subgroup analyses demonstrated that discrepancies involving a positive device output commonly occurred in the setting of recent intracranial surgery and intracranial hyperdense (non-hemorrhagic) lesions.

**Conclusion:** A PACS-integrated LLM approach, together with scalable frameworks for series selection and label generation, can provide radiologists with local insights about AI-enabled device performance. These insights may help radiologists interpret device outputs in clinical practice, particularly in scenarios associated with greater device-report discordance.

## Introduction

Artificial intelligence (AI)-enabled medical devices have demonstrated high levels of accuracy for many applications in radiology, leading to regulatory authorization and deployment into clinical practice.^1–3^ However, device performance may vary across institutions^4–7^ because of differences in disease prevalence,^8^ patient demographics,^9, 10^ and image acquisition protocols.^11^ Local evaluation, including pre-deployment baseline testing and ongoing monitoring post-deployment, can provide clinicians with important insights about device performance in their environment (Figure 1A).^12, 13^ This need is increasingly recognized by professional organizations including the American College of Radiology and is reflected in broader frameworks proposed in the peer-reviewed literature.^14–16^

**Figure 1A.**
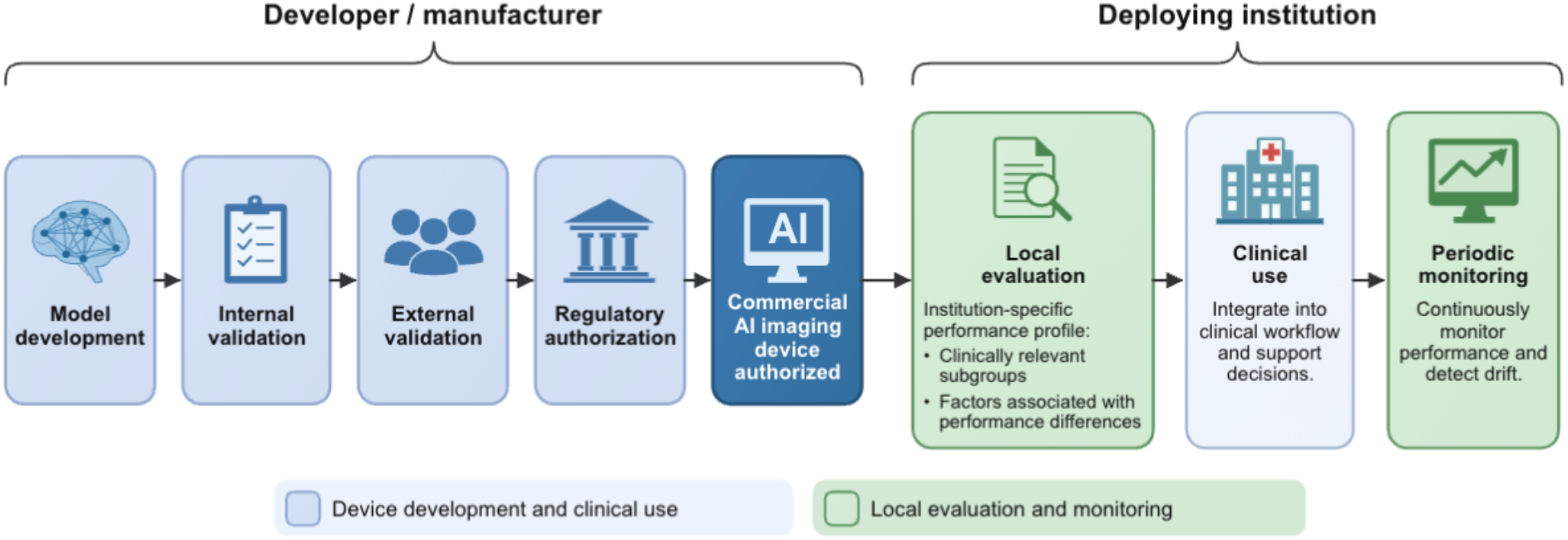
Key lifecycle components of a commercial Al imaging device and the role of local evaluation.

**Figure 1B.**
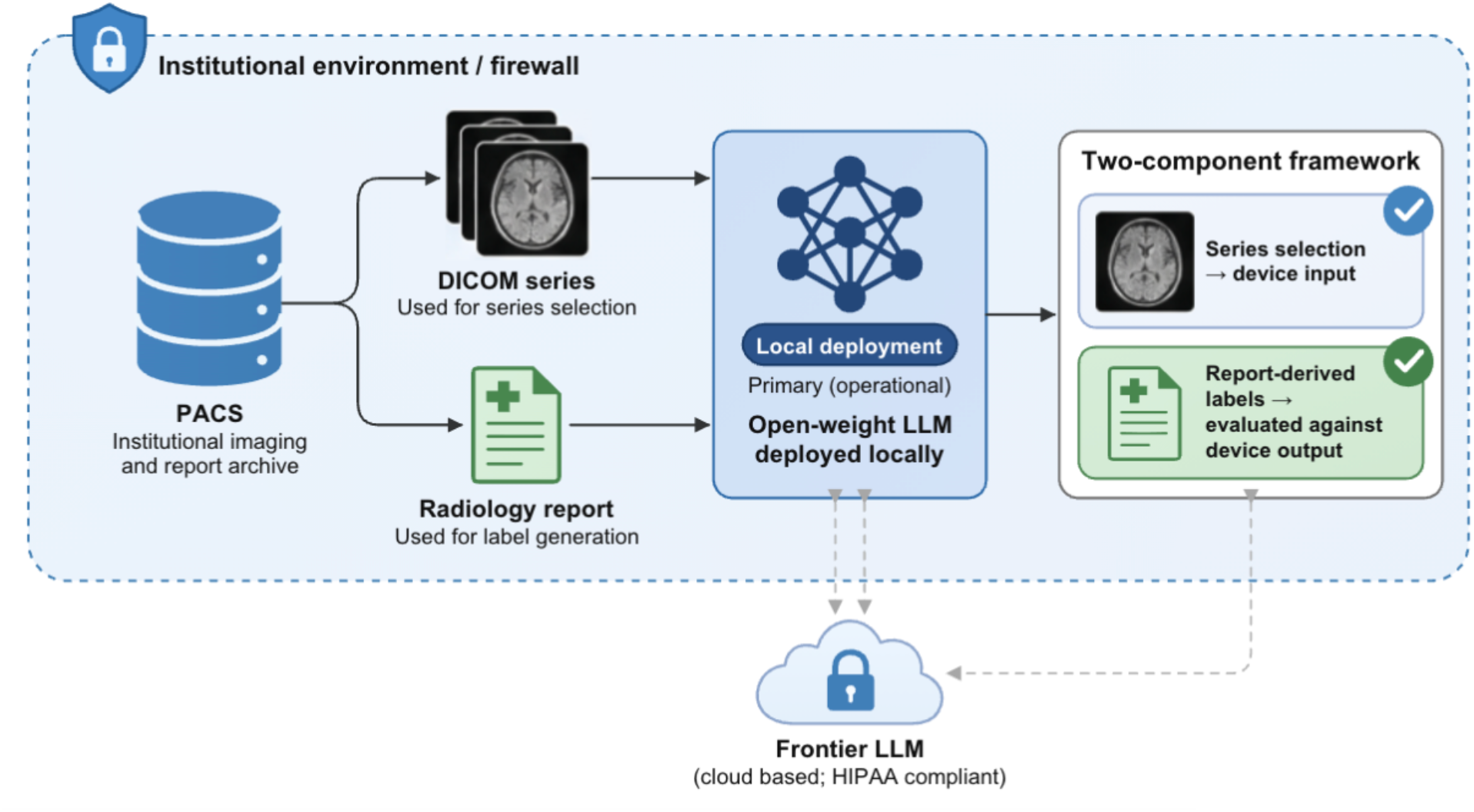
PACS-integrated LLM infrastructure for local evaluation.

Performing such evaluation at scale, however, is limited by technical, operational, and data-governance barriers.^17^ A single imaging examination yields multiple series, from which the appropriate series must be selected; many validation studies assume standardized protocols or rely on manual series curation.^18^ An appropriate comparator for the AI-enabled medical device is also necessary. Manual radiologist review of reports or images can provide a comparator but is labor-intensive and difficult to scale.^19^ Large language model (LLM) extraction offers a scalable alternative but introduces additional considerations, especially when there is discordance between device outputs and report-derived labels.^20^ Such discordance may reflect device error, label extraction error, or differences between device detection criteria and clinical reporting practices. Cloud-based LLM deployment may also raise data-governance and security concerns within institutional workflows.^21, 22^

To address these barriers, we integrated an LLM-based evaluation workflow with our picture archiving and communication system (PACS) environment (Figure 1B). It included an open-weight model deployed locally and a frontier model accessed through a Health Insurance Portability and Accountability Act (HIPAA)-compliant cloud environment. Using this infrastructure, we developed two scalable frameworks to automate key processes for the local evaluation of a commercial AI-enabled imaging device: identifying a candidate imaging series for device inference and generating report-derived labels using an LLM-as-a-judge approach without manual annotation.

We applied these frameworks to Purple AI Hyper Insight-ICH, a commercial device for intracranial hemorrhage (ICH) identification with US Food and Drug Administration (FDA) clearance.^23^ It had previously been shown to have a sensitivity of 94.4% and a specificity of 98.2% in an external validation set with almost 50,000 patients.^24^ Both frameworks were initially implemented using the open-weight model given it may be a preferred option for institutions due to data-governance requirements.^25^ Final report-derived label generation was additionally performed using the frontier model to assess the impact of LLM model choice on label consistency. We also performed subgroup analyses to identify clinical scenarios in which discrepancies between radiologist interpretations and device outputs may be more likely to occur.

## Methods

### Study design

This retrospective standalone model performance study was conducted using radiology cases from two hospitals within the Mass General Brigham (MGB) network between January 1, 2024, and June 30, 2025. One hospital was an academic medical center, while the other was a community hospital. The study was approved by the MGB Institutional Review Board with waiver of informed consent. It was conducted in accordance with relevant guidelines and regulations including HIPAA. This manuscript follows the Standards for Reporting Diagnostic Accuracy (STARD 2015) reporting guideline.

### Technical infrastructure

The LLMs used for this project were gpt-oss-120b, a local open-weight LLM within the institutional computing environment, and GPT-5.2, a frontier LLM accessed through a HIPAA-compliant cloud environment. They were integrated with the institutional PACS to support retrieval and processing of imaging metadata and radiology reports. Connections were established with the PACS report database such that the reports could be incorporated into prompts sent to LLMs. gpt-oss-120b was hosted on an H100 GPU with 80 GB memory. GPT-5.2 was accessed through an institutionally approved connection to the Azure AI Foundry. In addition to sending radiology reports, it was possible for external data to be uploaded in a comma separated value (CSV) file for the LLMs to process, which was used for DICOM metadata that had been manually extracted from imaging. LLM inputs and outputs were formatted as JSON to maintain structured fields.

### Study cohort

The cohort comprised 4,000 examinations with 2,000 selected in a consecutive manner from each hospital. The case-level inclusion criteria required head CT cases from adults aged 18 years or older and from unique patients; for patients with multiple examinations, only the earliest examination was retained. No additional exclusion criteria were applied. The series-level inclusion criteria, which were only applied once the 4,000 examinations were chosen, included axial plane orientation, 2mm to 5mm (inclusive) slice thickness, soft or standard kernel (excluding bone kernel), non-contrast, primary acquisition only (excluding reformatted images) and brain anatomic region.

### Series selection using LLM

For each examination, the locally deployed gpt-oss-120b model evaluated DICOM metadata for all series present within the studies for the six series-level inclusion criteria (see Series Selection Prompt in Supplemental Code). Each criterion was classified as green (eligible), yellow (indeterminate) or red (ineligible) based on the available metadata. The model performed this evaluation using nine Digital Imaging and Communications in Medicine (DICOM) attributes from each series: (0008,0008) Image Type, (0008,0070) Manufacturer, (0008,103E) Series Description, (0008,1090) Manufacturer’s Model Name, (0018,0050) Slice Thickness, (0018,1210) Convolution Kernel, (0020,0011) Series Number, (0028,1050) Window Center and (0028,1051) Window Width. These DICOM attributes were extracted based on the central slice of each series.

An overall series classification was assigned: green if all six criteria were satisfied, red if any criterion was not satisfied, or yellow if at least one criterion was indeterminate and none were ineligible. The model also generated a confidence score from 0 to 100, with scores of 100 for green series, 0 for red series, and intermediate scores for yellow series. For each examination, a single green series was selected when available and randomly selected if multiple were available; otherwise, the yellow series with the highest confidence score was selected. Examinations containing only red series were excluded from device inference.

### Model inference

The selected series were provided to the Purple AI Hyper Insight-ICH device (version 2.1.8) for model inference. This version was similar to the FDA-cleared version of Purple AI Hyper Insight-ICH, which provides binary outputs for ICH presence.^23^ For the present study, it was also configured to provide ICH volume.

### Report-derived label generation using LLM

Report-derived labels were generated through three sequential LLM prompting stages, designed to progressively characterize findings documented in radiology reports without requiring manual image annotation. Each stage considered only the Findings and Impression sections of the radiology report.

### Initial binary classification prompt (Prompt 1)

The locally deployed gpt-oss-120b model classified ICH as present or absent (see Binary Classification Prompt in Supplemental Code). This prompt provided examples of hemorrhage subtypes (epidural hematoma, subdural hematoma, subarachnoid hemorrhage, intraparenchymal hemorrhage, intracerebral hemorrhage, intraventricular hemorrhage) and examples of non-hemorrhagic findings (e.g., arteriovenous malformation) to guide hemorrhage identification without requiring subtype classification. It did not distinguish hemorrhage based on acuity.

### Discordance characterization prompt (Prompt 2)

Examinations with discordant binary report-derived labels and device outputs underwent LLM-based qualitative characterization. The locally deployed gpt-oss-120b model generated a description limited to 30 words of what it believed to be the single most salient imaging characteristic in each report that might explain the discordance (see Discordance Prompts in Supplemental Code). These descriptions were subsequently clustered into categories for each of report-positive/device-negative and report-negative/device-positive examinations using an institutionally available chatbot version of GPT-4o (see Discordance Clustering Prompts in Supplemental Code).

### Comprehensive prompt (Prompt 3)

The discordance analysis informed a third prompt that extracted 28 structured fields from each radiology report (see Comprehensive Prompt in Supplemental Code). This prompt included fields about ICH presence (present, absent or equivocal), hemorrhage subtype, minimal hemorrhage (e.g., punctate or trace), acuity, stable or resolving nature, quantitative dimensions, recent intracranial surgery, artifacts, calcifications, hyperdense (non-hemorrhagic) lesions, and extra-axial (non-hemorrhagic) collections. The prompt incorporated explicit rules for diagnostic uncertainty and required qualitative descriptions to be supported by exact report text. This prompt was applied to the entire analytic cohort using both the locally deployed gpt-oss-120b model and the frontier GPT-5.2 model.

### Statistical analysis

The quantitative statistical analyses were performed in R (version 4.6.1). The analyses involved binary output agreement analyses:^26^ positive percent agreement (PPA) and negative percent agreement (NPA) were calculated as the proportions of report-positive and report-negative examinations, respectively, with concordant device outputs. Positive predictive value (PPV) and negative predictive value (NPV) were calculated as the proportions of device-positive and device-negative examinations, respectively, with concordant report-derived labels. Confidence intervals (CIs) were two-sided and were calculated using bootstrapped intervals with 2,000 resamples.

For quantitative agreement analyses, report-derived labels were generated using the initial binary classification prompt (Prompt 1) with gpt-oss-120b and the comprehensive prompt (Prompt 3) with both gpt-oss-120b and GPT-5.2. The comprehensive prompt distinguished cases as being positive, negative or equivocal for ICH, which were reduced to binary outputs using two methods: considering equivocal cases as ICH-positive or excluding equivocal cases. To better match the inclusion criteria for many

ICH model studies, an additional PPA analysis considered only report-positive cases that contained acute or acute-on-chronic intracranial hemorrhage that was not equivocal or minimal (i.e., not punctate or trace). NPA was not calculated for this analysis because an equivalent restriction was not applied to the report-negative cases.

The PPA and NPA were further evaluated with subgroup comparisons across hemorrhage characteristics, including subtype, minimal hemorrhage, acuity, stability or resolving nature, and size, as well as the presence of recent intracranial surgery, intracranial artifacts, intracranial calcifications, intracranial hyperdense (non-hemorrhagic) lesions, and extra-axial (non-hemorrhagic) collections. Additional subgroup analyses examined hospital site, demographic characteristics, and scanner manufacturer. Volume-based analyses were not conducted because few reports contained quantitative hemorrhage measurements.

## Results

### Cohort and series selection

This study involved 4,000 consecutive non-contrast head CT examinations from two hospitals: 2,000 from an academic medical center and 2,000 from a community hospital. Among the 4,000 examinations, four were not accessible for deidentification, leaving 3,996 examinations comprising 31,422 series (Figure 2). Each series was classified using a traffic-light scheme based on six criteria: green (eligible for all criteria), yellow (indeterminate for one or more criteria without any ineligible criteria), or red (ineligible for at least one criterion). This automated classification yielded 4,205 green series (13.4%), 174 yellow series (0.6%), and 27,043 red series (86.1%). For each examination, a single green series was selected when available; otherwise, the highest-scoring yellow series was selected based on an LLM-generated numeric confidence score. A series was selected in 3,976 of the 3,996 examinations (99.5%): green in 3,918 (98.5%) and yellow in 58 (1.5%). The remaining 20 examinations (0.5%) contained only red series and were therefore excluded.

**Figure 2.**
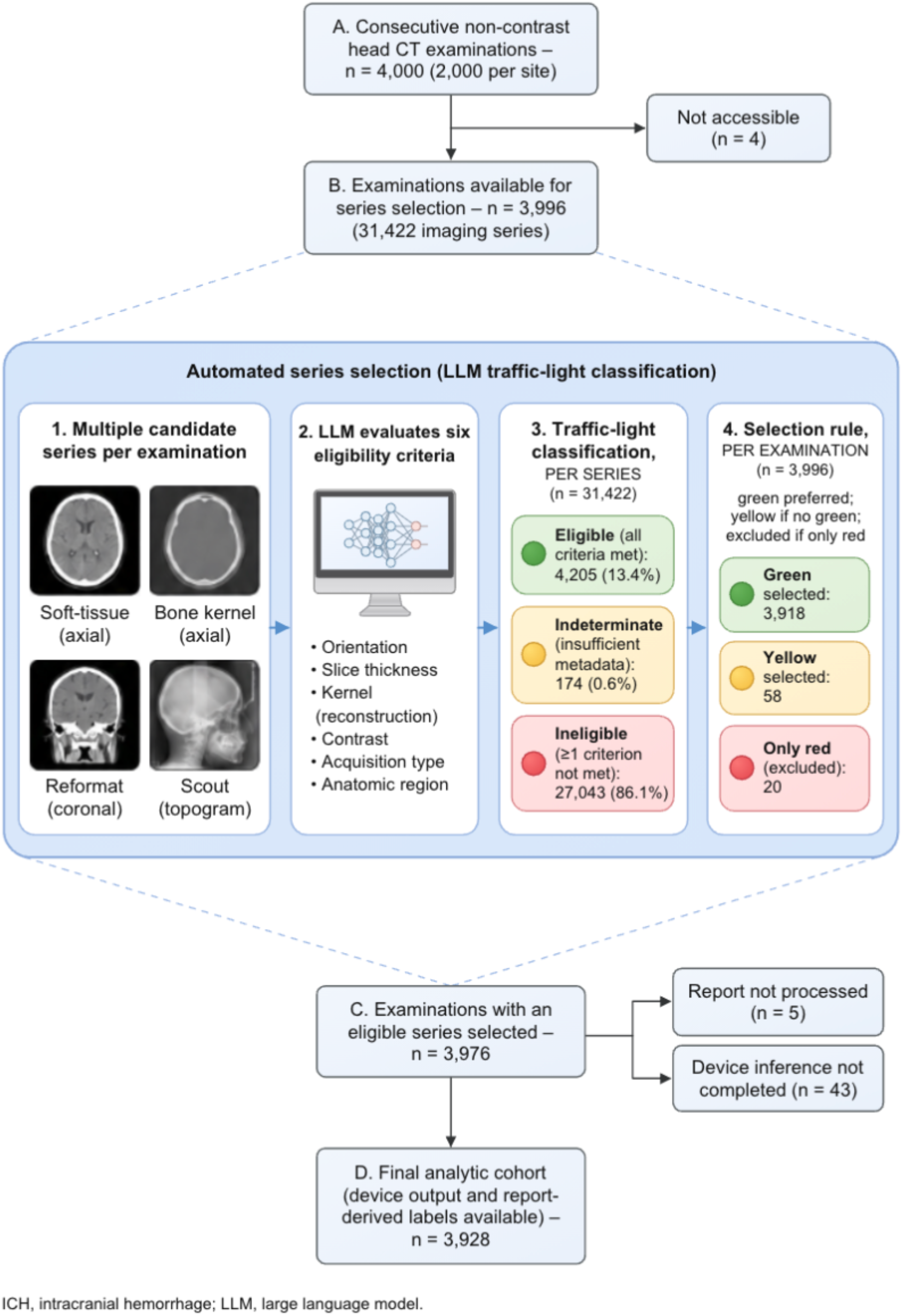
Cohort assembly and automated series selection.

Among these 3,976 examinations with a selected series, 48 examinations were excluded during downstream processing: five (0.1%) had inaccessible radiology reports and 43 (1.1%) did not complete device inference. The final cohort comprised 3,928 examinations with a mean patient age of 65.8 ± 20.0 years; 50.9% were from female patients (Table 1). The 43 examinations with unsuccessful inference included 38 from the 3,918 green series (1.0%) and five of 58 with yellow series (8.6%).

**Table 1:**
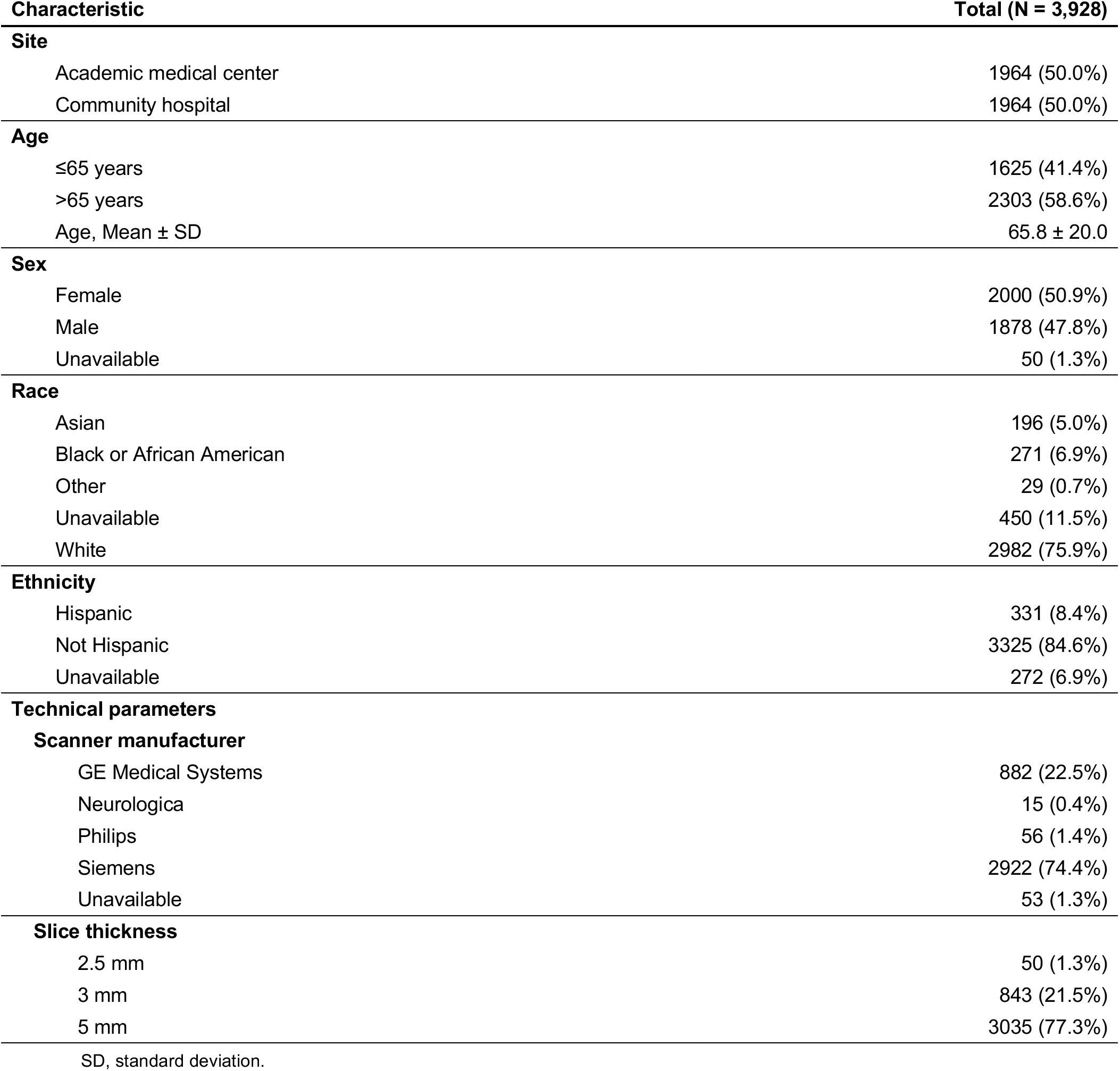
Cohort characteristics.

| **Characteristic** | **Total (N = 3,928)** |
|---|---|
| **Site** |  |
| Academic medical center | 1964 (50.0%) |
| Community hospital | 1964 (50.0%) |
| **Age** |  |
| ≤65 years | 1625 (41.4%) |
| >65 years | 2303 (58.6%) |
| Age, Mean ± SD | 65.8 ± 20.0 |
| **Sex** |  |
| Female | 2000 (50.9%) |
| Male | 1878 (47.8%) |
| Unavailable | 50 (1.3%) |
| **Race** |  |
| Asian | 196 (5.0%) |
| Black or African American | 271 (6.9%) |
| Other | 29 (0.7%) |
| Unavailable | 450 (11.5%) |
| White | 2982 (75.9%) |
| **Ethnicity** |  |
| Hispanic | 331 (8.4%) |
| Not Hispanic | 3325 (84.6%) |
| Unavailable | 272 (6.9%) |
| **Technical parameters** |  |
| **Scanner manufacturer** |  |
| GE Medical Systems | 882 (22.5%) |
| Neurologica | 15 (0.4%) |
| Philips | 56 (1.4%) |
| Siemens | 2922 (74.4%) |
| Unavailable | 53 (1.3%) |
| **Slice thickness** |  |
| 2.5 mm | 50 (1.3%) |
| 3 mm | 843 (21.5%) |
| 5 mm | 3035 (77.3%) |

SD, standard deviation.

### Report-derived label generation

Report-derived label generation was performed in three sequential stages. First, a locally deployed LLM generated a binary classification of ICH as present or absent from each radiology report (Prompt 1). Under this binary definition, ICH was present in 280 examinations (7.1%) and absent in 3,648 (92.9%). The initial LLM prompt did not distinguish hemorrhage acuity, while the device was designed specifically to detect acute ICH, creating anticipated discordance between report-derived labels and device output.^23^ The PPA was 64.3% (58.8 to 70.0%) and the negative percent agreement (NPA) was 94.6% (93.9 to 95.3%).

Second, reports from discordant examinations underwent LLM-based qualitative analysis (Prompt 2) to identify possible characteristics associated with discordance. These descriptions were then clustered using an LLM to identify recurrent patterns of discordance (Table 2). The most frequently identified characteristics among report-positive/device-negative cases included very small or trace hemorrhage; chronic, subacute or low attenuation; and equivocal report language. The most frequently identified characteristics among report-negative/device-positive cases included chronic parenchymal hypodensity; mass or vascular lesion; calcification or hyperdense vessel; and artifact, hardware or contrast staining. While this process identified recurrent patterns of discordance, some LLM-generated explanations were unsupported by broader clinical reasoning, highlighting the potential for hallucination in LLM analysis. For example, one case incorporated the reasoning “Chronic periventricular white matter hypodensities from small vessel disease may be misread as hemorrhage by the imaging model”.

**Table 2.** Characteristics associated with discordance between device output and report-derived labels.

| **Mechanism of discordance** | **n (%)** |
|---|---|
| **LLM positive, device negative discordant cases, n = 100** |  |
| *Imaging appearance* |  |
| Very small or trace hemorrhage | 45 (45.0) |
| Chronic, subacute or low attenuation | 22 (22.0) |
| Artifact or limited image quality | 4 (4.0) |
| *Subtotal* | *71 (71.0)* |
| *Report or label* |  |
| Equivocal report language | 20 (20.0) |
| Postoperative or residual, not active bleed | 9 (9.0) |
| *Subtotal* | *29 (29.0)* |
| *Not categorized* | *4 (4.0)* |
| **LLM negative, device positive discordant cases, n = 196** |  |
| *Imaging appearance* |  |
| Chronic parenchymal hypodensity | 36 (18.4) |
| Mass or vascular lesion | 24 (12.2) |
| Calcification or hyperdense vessel | 21 (10.7) |
| Artifact, hardware or contrast staining | 20 (10.2) |
| Postoperative or iatrogenic change | 17 (8.7) |
| Extracranial hematoma | 16 (8.2) |
| Extra-axial fluid collection | 14 (7.1) |
| Non-brain hyperdensity | 14 (7.1) |
| *Subtotal* | *162 (82.7)* |
| *Report or label* |  |
| Hemorrhage unmentioned or hedged in report | 33 (16.8) |
| *Not categorized* | *8 (4.1)* |

Discordant examinations were identified under the binary report-derived comparator. Percentages are of the discordant examinations in that group.
Categories were derived from structured analysis of discordant examinations and were intended to be mutually exclusive according to the LLM prompt; however, a small number of examinations were assigned to two categories. Category counts and percentages may therefore sum to more than the group total and 100%, respectively.

Third, the characteristics identified through the discordance analysis informed the development of a comprehensive prompt (Prompt 3), which extracted 28 structured report fields to characterize the examinations across the entire cohort. Prompt 3 was applied to both concordant and discordant examinations, allowing these characteristics to be evaluated systematically across the full cohort.

### Device agreement by case definition

Radiology reports frequently describe ICH with varying degrees of diagnostic uncertainty and clinical significance, creating multiple reasonable approaches to defining a positive examination for device evaluation. A key component incorporated into Prompt 3 was the creation of an “equivocal” category alongside “positive” and “negative” for the presence of ICH. PPA varied with the report-derived definition of a positive examination, decreasing to 59.9% when equivocal findings were classified as positive, and increasing to 66.4% when equivocal findings were excluded (Table 3). NPA was 94.7% (94.0 to 95.4%) with this prompt, while PPV was 49.2% (45.4 to 53.6%) and NPV was 96.5% (96.0 to 97.0%) when equivocal cases were considered positive.

**Table 3.** Device agreement by positive-case definition.

| **Positive-case definition** | **Positive N** | **Negative N** | **PPA (95% CI)** | **NPA (95% CI)** | **PPV (95% CI)** | **NPV (95% CI)** |
|---|---|---|---|---|---|---|
| Binary (Prompt 1): | 280 | 3648 | 64.3% (58.8–70.0) | 94.6% (93.9–95.3) | 47.9% (43.9–52.1) | 97.2% (96.7–97.6) |
| Equivocal included as positive (Prompt 3): | 309 | 3619 | 59.9% (54.2–65.2) | 94.7% (94.0–95.4) | 49.2% (45.4–53.6) | 96.5% (96.0–97.0) |
| Equivocal excluded (Prompt 3): | 256 | 3619 | 66.4% (60.5–72.2) | 94.7% (94.0–95.4) | 47.1% (43.1–51.2) | 97.6% (97.1–97.9) |
| Similar criteria for positive cases to published studies (Prompt 3): | 65 | — | 87.7% (79.2–95.0) | — | — | — |

PPA, positive percent agreement; NPA, negative percent agreement; PPV, positive predictive value; NPV, negative predictive value; CI, confidence interval.

The positive cases were also restricted based on other fields extracted by Prompt 3 to align more closely with common inclusion criteria for ICH detection model studies.

These criteria included non-equivocal hemorrhage, non-minimal hemorrhage (i.e., not punctate or trace), and acute or acute-on-chronic hemorrhage. PPA increased to 87.7% (79.2 to 95.0%) across 65 examinations.

### Device agreement by clinical subgroup

Subgroup analyses characterized the clinical and imaging contexts in which device outputs diverged from report-derived labels, indicating where radiologists using the device in the clinical environment may wish to apply additional scrutiny when interpreting device results. Among report-positive cases, PPA generally varied with hemorrhage characteristics associated with radiographic conspicuity (Table 4).

**Table 4.**
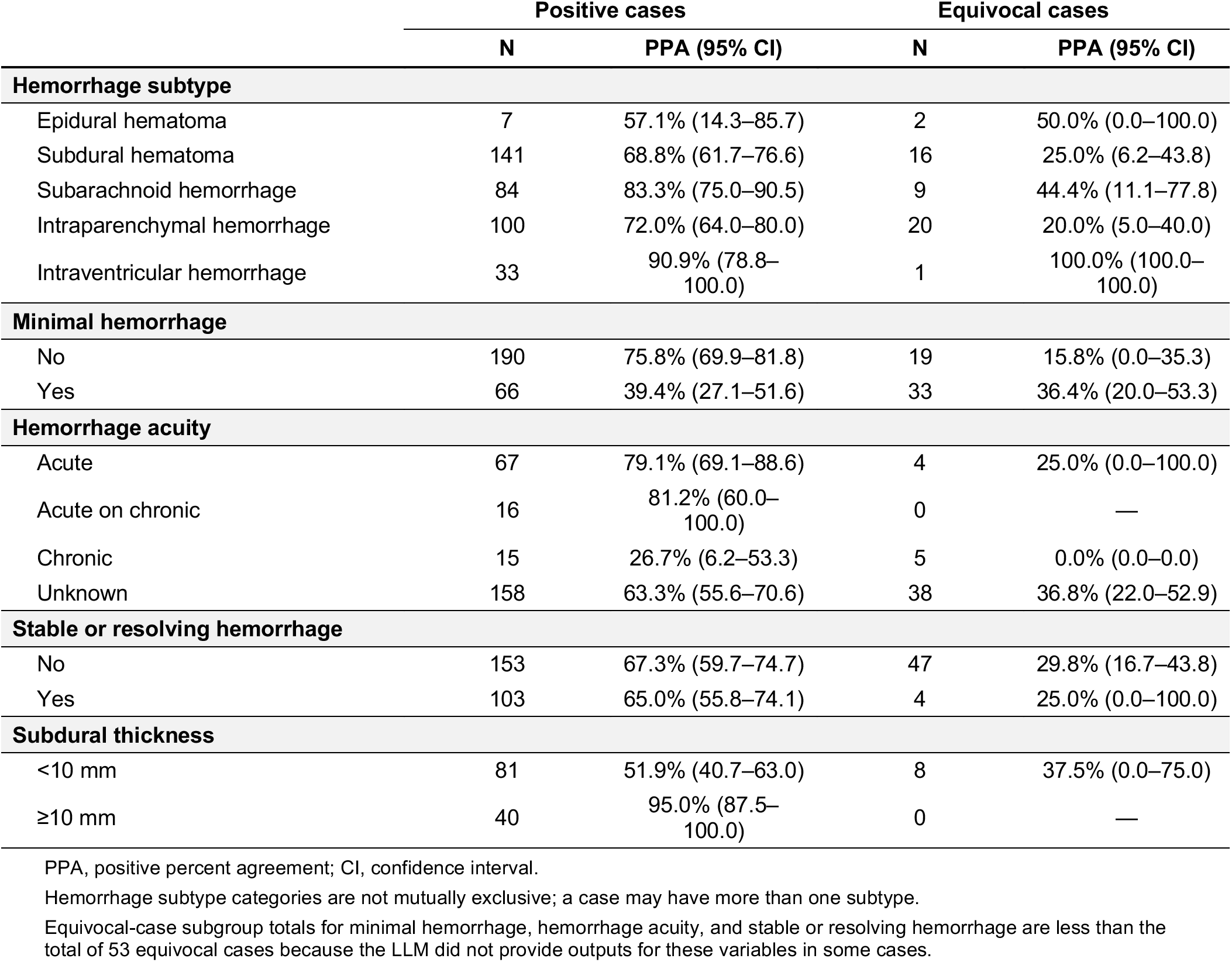
Device agreement in ICH-positive and equivocal cases, by clinical subgroup.

Agreement was substantially lower among minimal hemorrhages than among non-minimal hemorrhages (39.4% vs. 75.8%) and was markedly lower for chronic compared to acute hemorrhage (26.7% vs. 79.1%). Similarly, PPA increased with subdural hematoma thickness, from 51.9% for thickness <10 mm to 95.0% for collections ≥10 mm. Across hemorrhage subtypes, PPA ranged from 57.1% for epidural hematoma to 90.9% for intraventricular hemorrhage, with the most common subtypes, subdural hematoma and intraparenchymal hemorrhage, demonstrating PPAs of 68.8% and 72.0% respectively. Agreement was lower among equivocal cases across nearly all hemorrhage subtypes (subtype categories were not mutually exclusive, as individual examinations could contain multiple hemorrhage subtypes). PPA was similar regardless of whether hemorrhage was described as having a stable or resolving nature (65.0% vs. 67.3%).

Agreement also varied across structural and institutional subgroups (Table 5), with lower NPA observed among examinations with certain non-hemorrhagic imaging findings or recent intracranial surgery. NPA was 57.1% among cases with recent intracranial surgery compared with 95.0% among those without. Similarly, NPA was reduced in the presence of intracranial hyperdense non-hemorrhagic lesions (65.2% versus 95.1%) or extra-axial non-hemorrhagic collections (72.5% versus 95.3%). Smaller reductions were observed in the presence of intracranial artifact and calcification. Agreement also differed between the two participating hospitals, with higher PPA at the academic than community site (62.3% versus 52.1%).

**Table 5:** Device agreement across clinical, demographic, and technical subgroups.

|  | Equivocal included as positive |  |  |  | Equivocal excluded |  |  |  |
|---|---|---|---|---|---|---|---|---|
|  | Positive N | Negative N | PPA (95% CI) | NPA (95% CI) | Positive N | Negative N | PPA (95% CI) | NPA (95% CI) |
| **Recent intracranial surgery** |  |  |  |  |  |  |  |  |
| No | 266 | 3591 | 55.6% (49.6–61.7) | 95.0% (94.3–95.7) | 216 | 3591 | 62.5% (55.8–68.7) | 95.0% (94.3–95.7) |
| Yes | 43 | 28 | 86.0% (74.5–95.5) | 57.1% (38.5–76.0) | 40 | 28 | 87.5% (75.7–97.1) | 57.1% (38.5–75.9) |
| **Intracranial artifact** |  |  |  |  |  |  |  |  |
| Absent | 288 | 3432 | 60.8% (55.4–66.4) | 94.9% (94.2–95.6) | 242 | 3432 | 66.9% (60.8–72.8) | 94.9% (94.1–95.6) |
| Present | 21 | 187 | 47.6% (26.7–68.8) | 91.4% (87.4–95.6) | 14 | 187 | 57.1% (30.8–84.6) | 91.4% (87.2–95.2) |
| **Intracranial calcification** |  |  |  |  |  |  |  |  |
| Absent | 217 | 2494 | 62.7% (56.2–69.0) | 95.1% (94.2–95.9) | 186 | 2494 | 68.3% (61.5–74.5) | 95.1% (94.2–95.9) |
| Present | 92 | 1125 | 53.3% (43.2–64.0) | 94.0% (92.5–95.4) | 70 | 1125 | 61.4% (50.0–72.7) | 94.0% (92.6–95.3) |
| **Intracranial hyperdense (non-hemorrhagic) lesion** |  |  |  |  |  |  |  |  |
| Absent | 302 | 3573 | 60.3% (54.8–65.9) | 95.1% (94.4–95.8) | 252 | 3573 | 66.3% (60.3–72.1) | 95.1% (94.3–95.8) |
| Present | 7 | 46 | 42.9% (0.0–80.0) | 65.2% (51.0–79.1) | 4 | 46 | 75.0% (0.0–100.0) | 65.2% (51.0–78.3) |
| **Extra-axial (non-hemorrhagic) collection** |  |  |  |  |  |  |  |  |
| Absent | 279 | 3528 | 60.9% (55.1–66.7) | 95.3% (94.6–96.0) | 232 | 3528 | 67.2% (61.5–73.1) | 95.3% (94.6–96.0) |
| Present | 30 | 91 | 50.0% (32.3–68.8) | 72.5% (63.2–81.5) | 24 | 91 | 58.3% (38.1–78.3) | 72.5% (63.5–81.3) |
| **Hospital site** |  |  |  |  |  |  |  |  |
| Academic medical center | 236 | 1728 | 62.3% (56.2–68.5) | 94.3% (93.2–95.3) | 201 | 1728 | 67.7% (61.0–74.0) | 94.3% (93.2–95.3) |
| Community hospital | 73 | 1891 | 52.1% (41.0–64.3) | 95.1% (94.1–96.1) | 55 | 1891 | 61.8% (48.4–74.5) | 95.1% (94.1–96.1) |
| **Sex** |  |  |  |  |  |  |  |  |
| Female | 113 | 1887 | 67.3% (58.1–75.6) | 95.5% (94.5–96.5) | 93 | 1887 | 76.3% (67.4–84.5) | 95.5% (94.6–96.5) |
| Male | 194 | 1684 | 55.7% (48.9–62.7) | 93.8% (92.7–95.0) | 161 | 1684 | 60.9% (53.3–68.2) | 93.8% (92.6–94.9) |
| Unavailable | 2 | 48 | 50.0% (0.0–100.0) | 93.8% (86.0–100.0) | 2 | 48 | 50.0% (0.0–100.0) | 93.8% (86.0–100.0) |
| **Age** |  |  |  |  |  |  |  |  |
| ≤65 years | 119 | 1506 | 59.7% (50.9–68.2) | 95.3% (94.2–96.3) | 93 | 1506 | 65.6% (56.3–75.0) | 95.3% (94.2–96.3) |
| >65 years | 190 | 2113 | 60.0% (52.9–66.8) | 94.3% (93.3–95.3) | 163 | 2113 | 66.9% (59.5–73.8) | 94.3% (93.3–95.2) |
| **Race** |  |  |  |  |  |  |  |  |
| Asian | 25 | 171 | 60.0% (41.2–80.0) | 95.9% (92.7–98.8) | 23 | 171 | 60.9% (40.7–81.0) | 95.9% (92.6–98.8) |
| Black or African American | 21 | 250 | 52.4% (30.4–73.9) | 94.4% (91.4–97.2) | 18 | 250 | 61.1% (37.5–84.0) | 94.4% (91.5–97.2) |
| Other | 4 | 25 | 25.0% (0.0–100.0) | 88.0% (75.0–100.0) | 1 | 25 | 0.0% (0.0–0.0) | 88.0% (73.1–100.0) |
| Unavailable | 35 | 415 | 57.1% (39.4–74.2) | 94.2% (91.9–96.4) | 24 | 415 | 70.8% (50.0–88.0) | 94.2% (91.7–96.4) |
| White | 224 | 2758 | 61.6% (55.3–67.8) | 94.8% (94.0–95.6) | 190 | 2758 | 67.4% (61.0–73.9) | 94.8% (94.0–95.6) |
| **Ethnicity** |  |  |  |  |  |  |  |  |
| Hispanic | 28 | 303 | 60.7% (42.3–78.8) | 95.7% (93.0–98.0) | 20 | 303 | 70.0% (50.0–90.0) | 95.7% (93.2–97.7) |
| Not Hispanic | 254 | 3071 | 59.1% (52.9–65.0) | 94.8% (94.0–95.6) | 214 | 3071 | 64.5% (58.1–70.5) | 94.8% (94.1–95.6) |
| Unavailable | 27 | 245 | 66.7% (48.5–84.6) | 92.2% (88.8–95.4) | 22 | 245 | 81.8% (65.2–95.7) | 92.2% (88.7–95.4) |
| **Scanner manufacturer** |  |  |  |  |  |  |  |  |
| GE Medical Systems | 88 | 794 | 58.0% (48.2–68.4) | 96.1% (94.7–97.4) | 75 | 794 | 65.3% (54.5–75.9) | 96.1% (94.7–97.5) |
| Neurologica | 4 | 11 | 75.0% (0.0–100.0) | 72.7% (44.4–100.0) | 2 | 11 | 50.0% (0.0–100.0) | 72.7% (45.5–100.0) |
| Philips | 10 | 46 | 30.0% (0.0–62.5) | 95.7% (89.1–100.0) | 9 | 46 | 33.3% (0.0–66.7) | 95.7% (89.6–100.0) |
| Siemens | 204 | 2718 | 61.8% (54.9–68.3) | 94.4% (93.6–95.2) | 167 | 2718 | 68.9% (61.5–75.8) | 94.4% (93.6–95.2) |
| Unavailable | 3 | 50 | 66.7% (0.0–100.0) | 94.0% (86.3–100.0) | 3 | 50 | 66.7% (0.0–100.0) | 94.0% (86.3–100.0) |

PPA, positive percent agreement; NPA, negative percent agreement; CI, confidence interval.

### Device agreement by model choice

To evaluate the consistency of structured report-derived labels across LLMs, Prompt 3 was independently applied using the locally deployed gpt-oss-120b model and the cloud-based GPT-5.2 model. Report-derived classifications were highly concordant between the local and cloud models, agreeing in 3,884 of 3,928 examinations (98.9%). Among the 44 discordant examinations, 36 involved a change to or from an equivocal classification, while only 8 examinations changed directly between positive and negative. Device-report agreement estimates were similar using labels generated by either LLM across all positive-case definitions (Table 6).

**Table 6:** Device agreement by positive-case definition: local and frontier language model comparison.

| Positive-case definition | Positive N |  | Negative N |  | PPA (95% CI) |  | NPA (95% CI) |  | PPV (95% CI) |  | NPV (95% CI) |  |
|---|---|---|---|---|---|---|---|---|---|---|---|---|
|  | Local | gpt-5.2 | Local | gpt-5.2 | Local | gpt-5.2 | Local | gpt-5.2 | Local | gpt-5.2 | Local | gpt-5.2 |
| Binary (Prompt 1): | 280 | — | 3648 | — | 64.3% (58.8–70.0) | — | 94.6% (93.9–95.3) | — | 47.9% (43.9–52.1) | — | 97.2% (96.7–97.6) | — |
| Equivocal included as positive (Prompt 3): | 309 | 319 | 3619 | 3609 | 59.9% (54.2–65.2) | 59.6% (54.1–65.1) | 94.7% (94.0–95.4) | 94.8% (94.1–95.6) | 49.2% (45.4–53.6) | 50.5% (46.6–54.8) | 96.5% (96.0–97.0) | 96.4% (95.9–96.9) |
| Equivocal excluded (Prompt 3): | 256 | 238 | 3619 | 3609 | 66.4% (60.5–72.2) | 68.9% (63.1–74.6) | 94.7% (94.0–95.4) | 94.8% (94.1–95.5) | 47.1% (43.1–51.2) | 46.9% (42.8–51.0) | 97.6% (97.1–97.9) | 97.9% (97.5–98.3) |
| Similar criteria for positive cases to published studies (Prompt 3): | 65 | 54 | — | — | 87.7% (79.2–95.0) | 83.3% (72.1–92.5) | — | — | — | — | — | — |

PPA, positive percent agreement; NPA, negative percent agreement; PPV, positive predictive value; NPV, negative predictive value; CI, confidence interval.
Binary (Prompt 1) was evaluated only for the local model; the gpt-5.2 comparison was performed for Prompt 3 positive-case definitions only.
Positive N and Negative N differ between local and gpt-5.2 because report-derived labels were generated independently by each model.

## Discussion

This study presents the integration of LLMs with a PACS followed by their use in scalable frameworks for routine local evaluation of a commercial AI-enabled imaging device. Implementing such evaluation at scale commonly requires two processes: selecting the imaging series appropriate for device inference and generating a report-derived comparator from radiology reports. Applied to a commercial ICH detection device across 3,928 consecutive examinations at two hospitals, this study showed how these frameworks enabled automated evaluation using routinely generated data without manual annotation. By characterizing where device output and report-derived labels diverged, the frameworks also identified clinical scenarios in which radiologists may expect discrepancies between their interpretations and the device outputs.

The first framework addressed automated series selection. While series selection is less relevant during prospective clinical use because the acquisition parameters can be configured to align with an AI-enabled device, retrospective evaluations may require manual curation and limit scalability. We addressed this challenge using an LLM-based traffic-light scheme that classified each series as eligible (green), potentially eligible (yellow), or ineligible (red) against multiple DICOM-based criteria before selecting a single series for inference. Across 3,996 examinations comprising 31,422 imaging series, a series was selected for inference in 3,976 (99.5%) examinations, demonstrating that this prerequisite step could be automated at scale. Device inference failures were more frequent among yellow than green series (8.6% versus 1.0%), consistent with the expected greater uncertainty associated with yellow series. The use of DICOM-based criteria reflects that eligibility of series is still commonly established using discrete fields. It is possible that LLMs alongside vision-language models (VLMs) may enable series selection more directly from images moving forward.

The second framework addressed generation of the report-derived comparator. Rather than defining a comprehensive extraction schema at the outset, we developed it in three stages. Initial binary extraction established baseline agreement between the report-derived labels and the device outputs including identifying examinations with disagreement. Those discordant examinations were then analyzed using LLM-based qualitative characterization to identify potential imaging characteristics and reporting patterns associated with discordance. The characteristics identified during that analysis informed the fields subsequently extracted by a comprehensive prompt with 28 fields across the entire cohort. The objective was not to resolve discordance, but to learn from it, which differs from previous approaches that have generally used discordance as an evaluation outcome.^27, 28^ Our framework used discordant examinations to identify recurrent characteristics that could then be systematically evaluated across both concordant and discordant examinations, providing a local profile of device-report agreement that radiologists can draw on when using the device.

The iterative strategy demonstrated that agreement was influenced by both the definition of the report-derived comparator and the clinical context in which disagreement occurred. PPA was 64.3% under the binary comparator. Treating equivocal findings as positive lowered the PPA to 59.9%, whereas excluding them increased agreement to 66.4%. Aligning the definition with common inclusion criteria for many published studies further increased agreement to 87.7%; this agreement was also more consistent with the previously reported performance of the device.^24^ Such results are important for radiologists to be aware of as they work alongside AI-enabled devices in the clinical environment. One of the key changes in the definition was using only acute and acute-on-chronic hemorrhages, which aligns with the detection of acute ICH in the FDA 510(k) summary for this device.^23^ A consequential insight for radiologists to incorporate into clinical practice is that such a device may not identify chronic ICH (PPA 26.7% for chronic versus 79.1% for acute hemorrhage). It is possible that many other ICH devices would have a similar issue.

Radiology reports frequently describe findings in terms that do not reduce cleanly to the binary options of present or absent. These examinations are often excluded from model performance studies: one large-scale study had 10.35% positive examinations, yet excluded 6.21% of examinations that were post-operative and a further 1.70% that were indeterminate for ICH; the inclusion of these cases could have substantially changed the sensitivity.^27^ The current study deliberately retained all consecutive cases with cases only excluded for technical reasons (e.g., the case did not contain the appropriate series or inference was not completed). This approach was deliberate in order to consider all examinations that the model would encounter when used in the workflow.

This situation reflected the broader objective of local evaluation to characterize how device outputs align with radiologist interpretations within a specific clinical environment. A PPV of 47.9% provides a particularly interesting clinical perspective: in this cohort, approximately one in two positive device outputs did not correspond to a positive report-derived label, informing radiologists how frequently an alert may not align with their own report.

The principal strength of this study is that it demonstrates how a PACS-integrated LLM can enable scalable local evaluation. Here it was used on a retrospective cohort, representing the process for a pre-deployment assessment. It could be used in a similar way to support ongoing institutional monitoring of a device. Such assessments enable institutions to assess devices before adoption, after software updates, and periodically during clinical deployment as imaging protocols and workflows potentially evolve.

Although this study focused on ICH detection, the frameworks are intended to be adaptable across AI-enabled imaging applications. In addition, similar device-report agreement estimates were observed using report-derived labels generated by both a locally deployed open-weight model and frontier model accessed through a cloud environment, suggesting that the agreement findings were not substantially affected by model choice.

A key limitation of this study is that this PACS-integrated LLM was deployed at a single institution, albeit an institution that allowed both an academic medical center and community hospital to be assessed. The frameworks were also only applied to a single commercial device. Evaluation of the frameworks across additional devices and institutions will be needed to establish broader generalizability. Given a direct connection between the DICOM attributes and the LLMs was still being configured at the time of this study, this study also required manual extraction of the relevant DICOM fields from individual series before they were provided in a spreadsheet format to the PACS-integrated LLM. Our implementation additionally processed examinations individually and required substantial computational resources. At larger scale, components with established logic could instead be implemented using simpler deterministic tools, reserving LLM inference for tasks requiring flexible interpretation.

AI-enabled imaging devices continue to be adopted within radiology and knowledge of their performance at each institution is critical. This study demonstrated the integration of LLMs into a PACS and frameworks for using them to assess the agreement between an AI-enabled imaging device and existing radiology reports. This knowledge can provide radiologists with insights that allow them to practice more synergistically alongside these devices and hopefully deliver better care.

## Supporting information

STARD Checklist

## Data Availability

The test dataset generated for this study contains protected patient information. Some data may be available for research purposes from the corresponding author upon reasonable request.

