## Supplementary material for "Scalable frameworks for local evaluation of AI-enabled imaging devices using PACS-integrated large language models": Code supplement

**Series Selection Prompt**

*Please note that the specific DICOM attributes used as part of this prompt may vary based on institutional approaches for DICOM attributes. One item recognized following model inference for this project was that the 0008,0008 tag should not have been used for orientation given the DICOM standard stipulates that “AXIAL” means a “CT Cross-sectional image” (i.e., as opposed to a localizer image); this project also used the 0008,103E tag that commonly distinguished between orientations.*

@Model: openai/gpt-oss-120b@@

@MaxOutputTokens: 128000@@

@Reasoning:high@@

@FormatAsJson:@@

Prompt:

You are evaluating DICOM tags from individual series from head CTs. The overall purpose is to determine whether each series fulfills eligibility criteria for that series to be interpreted by an AI model.

This prompt includes the overall process in the Process Summary Section, descriptions of the criteria in the Background Context Section and specific output instructions in the Field Data Extraction Section. The input format is structured as a JSON string containing 10 fields (keys) that reflect the series UID and 9 other DICOM tags for each series. The output format is a JSON file containing exactly 16 fields (keys) as detailed in the Field Data Extraction Section. Each JSON object will show the responses based on the DICOM tags provided for that series. There will only be a single JSON object for each series. Please ensure that each output is valid JSON, double quotation marks are used for all JSON keys and string values, double quotation marks are NOT used within JSON string values, there is no commentary outside the JSON object and there are no trailing commas. Please note that text matching for inputs should be case insensitive and please ensure all outputs are in lower case. Please also limit JSON string responses to 30 words.

Process Summary Section:

The overall process is summarized as follows:

1.	Extract descriptions from the DICOM tags for each of six criteria.

2.	Based on the extracted description for each criterion, evaluate whether the series meets the respective eligibility requirement.

3.	Based on the eligibilities of all six criteria, evaluate whether the series meets the overall eligibility requirements.

For each of the six criteria, you will initially be asked to extract a description from the DICOM tags for them. You should then evaluate that description against the eligibility requirements for that respective criterion and use a ‘traffic light system’ to describe its eligibility. The traffic light system involves providing options of green (definitely fulfills criterion), red (definitely does not fulfill criterion) and yellow (insufficient information to determine if it fulfills or does not fulfill criterion). Please consider each criterion independently. Each criterion must be evaluated solely based on the explicit descriptions provided in the DICOM tags without inferring any information from other criteria or prior evaluations. DO NOT assume green or red if the relevant information is not provided in the DICOM tags; the output should be yellow if the eligibility cannot be definitively determined.

Once you have evaluated the six criteria, you will be asked to provide three outputs for the eligibility of the series overall. Firstly, you will use the ‘traffic light system’ for the eligibility of the series. This output should be green if the eligibilities for all criteria are green; red if any eligibilities are red; and yellow if no eligibilities are red and at least one is yellow. Secondly, you will provide an overall score between 0 and 100 to indicate your confidence that a series fulfills the eligibility criteria: red series should have a score of 0, yellow series should have a score between 1 and 99 based on your confidence (i.e., a score closer to 99 indicates greater confidence), green series should have a score of 100. Thirdly, you will provide a justification for your score.

Background Context Section:

The series eligibility criteria for this AI model are as follow:

1.	Orientation: axial plane

2.	Slice thickness: 2mm to 5mm (inclusive)

3.	Kernel: soft or standard (exclude bone kernel)

4.	Presence of contrast: non-contrast only

5.	Acquisition: primary acquisition only (exclude reformatted images)

6.	Anatomic region: brain

The file that you will receive provides the following nine DICOM tags in separate fields:

•	0008,0008 Image Type

•	0008,0070 Manufacturer

•	0008,103E Series Description

•	0008,1090 Manufacturer’s Model Name

•	0018,0050 Slice Thickness

•	0018,1210 Convolution Kernel

•	0020,0011 Series Number

•	0028,1050 Window Center

•	0028,1051 Window Width

The following background is provided for each eligibility criterion:

1. Orientation: information about the orientation is often included in the 0008,0008 and 0008,103E DICOM tags. The axial plane is commonly referred to with the terms “Axial” or “AX”. Other planes (which would indicate that the orientation is not in the axial plane) include the coronal plane (commonly referred to with the terms “Coronal” or “COR”) and the sagittal plane (commonly referred to with the terms “Sagittal” or “SAG”). This criterion should only consider orientation and should not consider other components of the acquisition (e.g., reformatted or derived acquisition, which is considered under the Acquisition criterion; or a topogram or scout image, which is considered under the Anatomic region criterion). If orientation terms across the 0008,0008 and 0008,103E DICOM tags conflict (e.g., “AX” and “COR” or “SAG”), do not assume a single orientation; instead classify the orientation as unknown and assign a traffic light classification of yellow.

2. Slice thickness: information about the slice thickness is usually included in the 0018,0050 DICOM tag, which provides a numeric value for the slice thickness in mm. The 0008,103E DICOM tag may also include the slice thickness either as a single numeric value (e.g., 2 or 5) or as a paired value with an “x” (e.g., 5x2.5) where the first value is the slice thickness. If the slice thickness is presented as paired values separated by a forward slash (e.g., 0.625/5), interpret the second value as the slice thickness and use it for evaluation; this format typically reflects acquisition versus reconstructed slice thickness, with the second value corresponding to the reconstructed slice thickness used for interpretation. Slice thickness must be evaluated as a numeric value and compared to the numeric values from the eligibility criteria above; a numeric value outside the range of eligibility above should result in a classification of “red” for this criterion.

3. Kernel: information about the kernel is usually included in the 0008,103E and 0018,1210 DICOM tags. The 0028,1050 and 0028,1051 DICOM tags may also show window center and window width values that are suggestive of the kernel but they should not override explicit kernel information contained in other tags. Common kernels include “soft”, “standard”, “bone”.

4. Contrast: information about contrast is usually included in the 0008,103E DICOM tag. A contrast series is commonly referred to with the terms “W”, “I+”, “C+”, “With”, “Post”. A non-contrast series is commonly referred to with the terms “WO”, “I-”, “C-”, “Without”, “Pre”.

5. Acquisition: information about the acquisition is usually included in the 0008,0008 and 0008,103E DICOM tags. A primary acquisition is commonly referred to with the term “primary”. A non-primary acquisition is commonly referred to with the terms “secondary”, “derived”, “reformatted”, “MPR”. If conflicting terms are present (e.g., “primary” and “MPR”), the series should be considered a non-primary acquisition as the terms likely reflect that the primary acquisition was subsequently processed to become a non-primary acquisition. The 0020,0011 DICOM tag may also suggest that a series is primary or non-primary but should not override explicit tags: the primary acquisition series commonly use consecutive series numbers that commence at “1”, while the non-primary acquisition series commonly use series numbers that start in the hundreds (e.g., “300” or “400”).

6. Anatomic region: information about the anatomic region is usually included in the 0008,103E DICOM tag. Common terms to indicate that the anatomic region is the brain include “br”, “brain”, “head”. Terms that indicate that the anatomic region is not the brain include “face”, “c-spine”, “spine”, “inner ear”, “craniofacial”. In addition, terms that suggest that the anatomic region has not been completely captured because the series is not intended for diagnostic purposes include “scout”, “topogram”, “monitoring”, “premonitoring”, “dose”, “surview”; the presence of these terms should result in a classification of “red” for this criterion.

Field Data Extraction Section:

This section describes the fields that should be included in the generated JSON file output.

Field 1, Title, “Series UID”: Output the series UID

Fields 2 to 13 include two fields for each of the six criteria as described in the Background Context Section. The first field is titled “[Criterion] qualitative” and the second field is titled “[Criterion] traffic light”. For the first field, output your description for that field (e.g., “axial” may be an output for Orientation and “2mm” may be an output for slice thickness); use “unknown” if you cannot determine a description (in which case the traffic light classification should be “yellow”). For the second field, output the traffic light classification (“green”, “yellow”, “red”) based on whether your outputted description in the first field fulfills the eligibility criteria described in the Background Context Section.

Fields 2-3, Criterion: Orientation.

Fields 4-5, Criterion: Slice thickness.

Fields 6-7, Criterion: Kernel.

Fields 8-9, Criterion: Presence of contrast.

Fields 10-11, Criterion: Acquisition.

Fields 12-13, Criterion: Anatomic region.

Fields 14 to 16 are three fields to describe the series overall.

Field 14, Title, “Overall classification”: Output the traffic light classification (“green”, “yellow”, “red”) for whether this series fulfills all six criteria.

Field 15, Title, “Overall score”: Output your score between 0 and 100 to indicate your confidence that a series fulfills the criteria.

Field 16, Title, “Overall score justification”: Output your justification for your overall score provided in Field 15. Cite the specific tag(s) and tag value(s) in providing this justification.

**Binary Classification Prompt**

@Model: gpt-5.2-latest@@ | @Model: openai/gpt-oss-120b@@

@MaxOutputTokens: 128000@@

// @Reasoning:high@@

@FormatAsJson:@@

Prompt:

You are evaluating radiology reports of head CTs and extracting key information to assess the presence of intracranial hemorrhage. This prompt includes the definitions in the Clinical Context Section and specific extraction instructions in the Field Data Extraction Section.

For each radiology report, please only consider the "Findings" and "Impression" sections of the @CurrentReport@@. DO NOT consider the @PriorReports@@. The output format is a JSON file containing exactly 2 fields (keys) as detailed in the Field Data Extraction Section. Each JSON object will show the extracted information from an individual report for a head CT. There will only be a single JSON object for each head CT. Please ensure that each output is valid JSON, double quotation marks are used for all JSON keys and string values, double quotation marks are NOT used within JSON string values, there is no commentary outside the JSON object and there are no trailing commas.

Clinical Context Section:

This section provides definitions that should be directly referenced as part of the Field Data Extraction.

Intracranial refers specifically to anything within the skull. This task focuses on intracranial findings and should only consider intracranial findings.

An intracranial hemorrhage is defined as any hemorrhage within the skull. It may consist of the following subtypes of hemorrhage: epidural hematoma, subdural hematoma, subarachnoid hemorrhage, intraparenchymal hemorrhage, intracerebral hemorrhage, intraventricular hemorrhage. An intracranial hemorrhage does not include arteriovenous malformation, cavernoma, developmental venous anomaly, capillary telangiectasia. Multiple types of intracranial hemorrhage may be seen in an individual report. The terms hemorrhage and hematoma may be used interchangeably. There are two possible outputs for the presence of intracranial hemorrhage: "present" or "absent".

Language indicating complete resolution of hemorrhage (e.g., "resolved", "no longer seen", "no residual hemorrhage") should be considered "absent". Language describing persistent hemorrhage (e.g., "stable", "unchanged", "redemonstrated") should be considered "present".

Petechial hemorrhage is a form of intraparenchymal hemorrhage and should be classified as "present". A collection (e.g., extra-axial collection) should only be considered "present" for intracranial hemorrhage if it is explicitly described as a hemorrhage or hematoma. Collections described as hygroma or fluid without hemorrhage should be considered "absent".

Field Data Extraction Section:

This section describes the fields that should be included in the generated JSON file output.

Field 1, Title, "Accession number": Output the accession number.

Field 2, Title, "Intracranial hemorrhage present?": Using the definitions in the Clinical Context Section as the sole basis for determination, output "Intracranial hemorrhage PRESENT" if intracranial hemorrhage is present; output "Intracranial hemorrhage ABSENT" if intracranial hemorrhage is absent.

**Discordance Prompts**

*Please note that the terms “false negative” and “false positive” were used within the prompt for simplicity, while recognizing the comparator was the LLM-derived binary classifications (i.e., not a reference standard produced directly by a radiologist).*

*False negative prompt*

@Model: openai/gpt-oss-120b@@

@MaxOutputTokens: 128000@@

@FormatAsJson:@@

Prompt:

You are evaluating radiology reports of head CTs where there was a discordance in hemorrhage identification between an LLM-based classification of the radiology report and a commercial AI imaging model’s assessment of the image itself. In false negative cases the LLM-based classification identified intracranial hemorrhage as present, but the imaging model identified intracranial hemorrhage as absent. The goal is to identify the single most salient imaging characteristic described in the report that may explain this discordance.

For each report, consider only the "Findings" and "Impression" sections of the @CurrentReport@@. DO NOT consider the @PriorReports@@.

The output format is a JSON file containing exactly 2 fields. Please ensure valid JSON, double quotation marks for all keys and string values, no double quotation marks within string values, no commentary outside the JSON object, and no trailing commas.

Field Data Extraction Section:

Field 1, "Accession number": Output the accession number.

Field 2, "Salient imaging characteristic": Provide a 1-2 sentence description of the single most salient imaging characteristic described in the report that may explain the discordance. Descriptions should be concise and consistent in style across cases as outputs will be used to identify common patterns across the full discordant case set. Limit to 30 words.

*False positive prompt*

@Model: openai/gpt-oss-120b@@

@MaxOutputTokens: 128000@@

@FormatAsJson:@@

Prompt:

You are evaluating radiology reports of head CTs where there was a discordance in hemorrhage identification between an LLM-based classification of the radiology report and a commercial AI imaging model’s assessment of the image itself. In false positive cases the imaging model identified intracranial hemorrhage as present, but the LLM-based classification identified intracranial hemorrhage as absent. The goal is to identify the single most salient imaging characteristic described in the report that may explain this discordance.

For each report, consider only the "Findings" and "Impression" sections of the @CurrentReport@@. DO NOT consider the @PriorReports@@.

The output format is a JSON file containing exactly 2 fields. Please ensure valid JSON, double quotation marks for all keys and string values, no double quotation marks within string values, no commentary outside the JSON object, and no trailing commas.

Field Data Extraction Section:

Field 1, "Accession number": Output the accession number.

Field 2, "Salient imaging characteristic": Provide a 1-2 sentence description of the single most salient imaging characteristic described in the report that may explain the discordance. Descriptions should be concise and consistent in style across cases as outputs will be used to identify common patterns across the full discordant case set. Limit to 30 words.

**Discordance Clustering Prompts**

*Please note that the terms “false negative” and “false positive” were used within the prompt for simplicity, while recognizing the comparator was the LLM-derived binary classifications (i.e., not a reference standard produced directly by a radiologist).*

*False negative prompt*

This analysis is part of a local institutional validation study of a commercial AI imaging model for intracranial hemorrhage detection on non-contrast head CT. The model was evaluated against a ground truth derived from an LLM-based classification of radiology reports.

A set of discordant cases has been identified where the LLM-based classification and the imaging model disagreed on the presence or absence of intracranial hemorrhage. In false negative cases the LLM-based classification identified intracranial hemorrhage as present but the imaging model identified intracranial hemorrhage as absent. For each discordant false negative case, a brief description of the single most salient imaging characteristic relevant to the discordance was generated directly from the radiology report text using a local LLM. These descriptions are listed below.

Please review all descriptions and identify the smallest set of parsimonious and mutually exclusive categories that captures the majority of cases. Do not create categories unless clearly supported by multiple cases. For each category provide a short name, a 1-2 sentence definition, the number of cases, and 2-3 representative examples.

*False positive prompt*

This analysis is part of a local institutional validation study of a commercial AI imaging model for intracranial hemorrhage detection on non-contrast head CT. The model was evaluated against a ground truth derived from an LLM-based classification of radiology reports.

A set of discordant cases has been identified where the LLM-based classification and the imaging model disagreed on the presence or absence of intracranial hemorrhage. In false negative cases the LLM-based classification identified intracranial hemorrhage as present but the imaging model identified intracranial hemorrhage as absent. In false positive cases the imaging model identified intracranial hemorrhage as present but the LLM-based classification identified intracranial hemorrhage as absent. For each discordant case a brief description of the single most salient imaging characteristic relevant to the discordance was generated directly from the radiology report text using a local LLM. These descriptions are listed below.

Please review all descriptions and identify the smallest set of parsimonious and mutually exclusive categories that captures the majority of cases. Do not create categories unless clearly supported by multiple cases.

For each category provide a short name, a 1-2 sentence definition, the number of cases, and 2-3 representative examples.

**Comprehensive Prompt**

@Model: openai/gpt-oss-120b@@
@MaxOutputTokens: 128000@@
@FormatAsJson:@@

Alternate for @Model: gpt-5.2-latest@@

@Model: gpt-5.2-latest@@
@MaxOutputTokens: 128000@@
@Reasoning:high@@
@FormatAsJson:@@

Prompt:
You are evaluating radiology reports of head CTs and extracting key information. This information includes the presence of intracranial hemorrhage and other characteristics as specified below. This prompt includes the overall process in the Process Summary Section, definitions in the Clinical Context Section and specific extraction instructions in the Field Data Extraction Section.

For each radiology report, please only consider the “Findings” and “Impression” sections of the @CurrentReport@@. DO NOT consider the @PriorReports@@. Each output should be directly referenced from specific text in the “Findings” and “Impression” sections. All qualitative and dimensional fields must contain exact phrases copied from the report. DO NOT paraphrase. For fields requiring subtype labeling, append the subtype in parentheses after the exact copied phrase. The output format is a JSON file containing exactly 28 fields (keys) as detailed in the Field Data Extraction Section. Each JSON object will show the extracted information from an individual report for a head CT. There will only be a single JSON object for each head CT. Please ensure that each output is valid JSON, double quotation marks are used for all JSON keys and string values, double quotation marks are NOT used within JSON string values, there is no commentary outside the JSON object and there are no trailing commas. Please also limit JSON string responses to 30 words.

Process Summary Section:
This section provides a summary of the overall process.
Step 1: determine the presence of intracranial hemorrhage (output in Field 2).
Step 2: determine the subtype(s) of intracranial hemorrhage (output in Fields 3 to 7).
Step 3: determine the characteristics of intracranial hemorrhage including acuity and dimensions (output in Fields 8 to 18).
Step 4: determine the presence of other findings and clinical context (output in Fields 19 to 28).

Clinical Context Section:
This section provides definitions that should be directly referenced as part of the Field Data Extraction.

Intracranial refers specifically to anything within the skull. This task focuses on intracranial findings and should only consider intracranial findings.

An intracranial hemorrhage is defined as any hemorrhage within the skull. It may consist of the following subtypes of hemorrhage: epidural hematoma, subdural hematoma, subarachnoid hemorrhage, intraparenchymal hemorrhage, intracerebral hemorrhage, intraventricular hemorrhage. An intracranial hemorrhage does not include arteriovenous malformation, cavernoma, developmental venous anomaly, capillary telangiectasia. Multiple types of intracranial hemorrhage may be seen in an individual report. The terms hemorrhage and hematoma may be used interchangeably. There are three possible outputs about the presence of intracranial hemorrhage: “present”, “equivocal”, “absent”. Language indicating a reasonable likelihood of hemorrhage (e.g., “likely”, “probable”, “suggestive of”) should be considered “present”. Language that explicitly suggests uncertainty (e.g., “equivocal”, “cannot exclude”, “possible”, “may represent”) should be considered “equivocal”. Hemorrhage that is presented as part of a differential diagnosis (e.g., “hemorrhage versus calcification”) without clear attribution should be considered “equivocal”. Language indicating complete resolution of hemorrhage (e.g., “resolved”, “no longer seen”, “no residual hemorrhage”) should be considered “absent”. Language describing persistent hemorrhage (e.g., “stable”, “unchanged”, “redemonstrated”) should be considered “present”.

The terms intraparenchymal and intracerebral refer to hemorrhage specifically within the brain tissue. These terms may be used interchangeably. Petechiae or petechial hemorrhage is a form of intraparenchymal hemorrhage. Chronic microhemorrhages or chronic microbleeds without acute blood products should be considered “absent” for this task.

An extra-axial collection refers to a collection of fluid in the extra-axial space, which includes the epidural, subdural and subarachnoid spaces. An extra-axial collection may include an intracranial hemorrhage (e.g., subdural hematoma). An extra-axial collection may also occur without an intracranial hemorrhage (e.g., subdural hygroma). A case should only be classified as “present” for intracranial hemorrhage if the extra-axial collection is explicitly described as hemorrhage or if there is an intracranial hemorrhage separate to the extra-axial collection. The term “extra-axial (non-hemorrhagic) collection” does not include hemorrhagic collections (which are captured by other extracted fields), although it is possible for a case that contains an intracranial hemorrhage to also contain an extra-axial (non-hemorrhagic) collection.

The acuity of a hemorrhage is commonly described as “acute”, “chronic”, “acute on chronic”. The last option (acute on chronic) is a hemorrhage that includes features of both acute and chronic hemorrhages. An acute hemorrhage may also be described as hyperdense. A chronic hemorrhage may also be described as isodense or hypodense.

The term “minimal hemorrhage” is a term for this task that describes an intracranial hemorrhage that has a very small volume using descriptors such as “minimal”, “trace”, “punctate”. Petechial hemorrhage is considered minimal hemorrhage. A study is classified as minimal hemorrhage only if all hemorrhage within the case is considered minimal; if any non-minimal hemorrhage is present, a case would not be considered to contain minimal hemorrhage.

The term “stable or resolving hemorrhage” refers to an intracranial hemorrhage for which the report captures that it is stable or resolved compared to another recent scan. The descriptors may include “decreased conspicuity”, “improving”, “interval reduction”.

Subdural and epidural hematomas may be measured with a maximal thickness or depth as the key dimension. Intraparenchymal hemorrhages may be measured with a volume, which reports may describe with a calculated volume or with dimensions that can then be used to estimate the volume with an ellipsoid formula [(length * width * thickness or height) / 2]. Each report may or may not provide the full set of measurements that would be required to calculate the volume of an intraparenchymal hemorrhage. DO NOT calculate hemorrhage volume unless all three orthogonal dimensions are explicitly provided.

This prompt also considers the presence of mimics and other findings.

Recent intracranial surgery is considered to have occurred within 7 days. When an exact time is not provided, it may be suggested by the presence of radiographic findings including pneumocephalus or surgical drain. Chronic postoperative changes without evidence of recent intervention should not be considered recent surgery.

Intracranial artifacts may include motion or metal artifacts including from dental implants and surgical clips. They should only be considered present if they are described as impacting the ability to visualize intracranial structures.

Intracranial hyperdense (non-hemorrhagic) lesion refers to a lesion that is considered hyperdense. It includes hyperdense tumors and hyperdense vascular malformations (e.g., arteriovenous malformations). It only includes lesions that are explicitly described as hyperdense or similar terms (e.g., “hyperattenuating”). It does not include hemorrhagic lesions (which are captured by other extracted fields), although it is possible for a case that contains an intracranial hemorrhage to also contain a hyperdense (non-hemorrhagic) lesion.

Field Data Extraction Section: 
This section describes the fields that should be included in the generated JSON file output.

Field 1, Title, “Accession number”: Output the accession number.

Field 2, Title, “Intracranial hemorrhage present?”: If an intracranial hemorrhage is present, output “Intracranial hemorrhage PRESENT”. If an intracranial hemorrhage is equivocal, output “Intracranial hemorrhage EQUIVOCAL”. If an intracranial hemorrhage is absent, output “Intracranial hemorrhage ABSENT”. For the downstream fields about the subtypes and characterization of the intracranial hemorrhage (Fields 3 to 19), please consider both present and equivocal hemorrhages.

Fields 3–7 represent subtype classifications of intracranial hemorrhage. If field 2 is “Intracranial hemorrhage PRESENT” or “Intracranial hemorrhage EQUIVOCAL”, for each of fields 3-7, output “YES [Subtype]” if the intracranial hemorrhage is described as present or equivocal for the respective subtype in the report and, else, output “NO [Subtype]”. If field 2 is “Intracranial hemorrhage ABSENT”, output “N/A” for each of fields 3-7.

Field 3: Title: “Epidural hematoma”, Subtype: Epidural hematoma.

Field 4, Title: “Subdural hematoma”, Subtype: Subdural hematoma.

Field 5, Title: “Subarachnoid hemorrhage”, Subtype: Subarachnoid hemorrhage.

Field 6, Title: “Intraparenchymal hemorrhage”, Subtype: Intraparenchymal hemorrhage.

Field 7, Title: “Intraventricular hemorrhage”, Subtype: Intraventricular hemorrhage.

Field 8, Title “Minimal hemorrhage?”: If field 2 is “Intracranial hemorrhage PRESENT” or “Intracranial hemorrhage EQUIVOCAL”, determine if the report suggests that this CT study only includes minimal hemorrhage amongst all subtypes of hemorrhage; if so, output “YES Minimal hemorrhage” and, else, output “NO Minimal hemorrhage”. If field 2 is “Intracranial hemorrhage ABSENT”, output “N/A”.

Field 9, Title, “Minimal hemorrhage qualitative”: If field 8 is “YES Minimal hemorrhage”, output the phrasing from the report that suggests the hemorrhage is minimal. If field 8 is “NO Minimal hemorrhage” or “N/A”, output “N/A”.

Field 10, Title, “Hemorrhage acuity”: If field 2 is “Intracranial hemorrhage PRESENT” or “Intracranial hemorrhage EQUIVOCAL”, determine and output the acuity of any present or equivocal hemorrhages. The four options to output include “Acute”, “Chronic”, “Acute on chronic”, “Unknown acuity”. Output acuity as a semicolon-separated list and include each hemorrhage subtype in parentheses (e.g., “Chronic (Subdural hematoma); Acute (Subarachnoid hemorrhage)”). If the report does not describe the acuity for a hemorrhage subtype, use “Unknown acuity” even if the acuity is known for other subtypes. Include one acuity entry for each hemorrhage subtype marked “YES [Subtype]” in fields 3-7. If field 2 is “Intracranial hemorrhage ABSENT”, output “N/A”.

Field 11, Title, “Hemorrhage acuity qualitative”: If field 10 includes acuities that incorporate “Acute”, “Chronic” or “Acute on chronic”, output the phrasing from the report that was used to determine the acuity (or multiple acuities). If field 10 is “N/A” or only incorporates “Unknown acuity”, output “N/A”.

Field 12, Title “Stable or resolving hemorrhage?”: If field 2 is “Intracranial hemorrhage PRESENT” or “Intracranial hemorrhage EQUIVOCAL”, determine if the report suggests that this CT study is a follow-up study that shows the hemorrhage is stable or resolving; if so, output “YES Stable or resolving hemorrhage” and, else, output “NO Stable or resolving hemorrhage”. If field 2 is “Intracranial hemorrhage ABSENT”, output “N/A”.

Field 13, Title, “Stable or resolving hemorrhage qualitative”: If field 12 is “YES Stable or resolving hemorrhage”, output the phrasing from the report that suggests the hemorrhage is stable or resolving. If field 12 is “NO Stable or resolving hemorrhage” or “N/A”, output “N/A”.

Field 14, Title “Hemorrhage quantitative dimensions listed?”: If field 2 is “Intracranial hemorrhage PRESENT” or “Intracranial hemorrhage EQUIVOCAL”, determine if there are numeric dimensions that directly measure the size of intracranial hemorrhage (e.g., length, width, height, thickness, volume); if so, output “YES Dimensions listed” and, else, output “NO Dimensions listed”. If field 2 is “Intracranial hemorrhage ABSENT”, output “N/A”.

Field 15, Title “Hemorrhage dimensions”: If field 14 is “YES Dimensions listed”, output the text from the report that describes the dimensions. As part of this output, also include in parentheses the type of intracranial hemorrhage for which these dimensions apply. If there are multiple hemorrhages with associated dimensions, list the dimensions for each hemorrhage in the same JSON string value as a semicolon-separated list. If field 14 is “NO Dimensions listed” or “N/A”, output “N/A”.

Field 16, Title “Subdural or epidural hematoma thickness”: If field 14 is “YES Dimensions listed” and the dimensions include subdural hematoma or epidural hematoma thickness, output the numeric value for the thickness in mm. If thickness is provided with different units (e.g., cm) then convert to mm. If there are multiple subdural hematoma or epidural hematoma thicknesses provided, provide only the maximum thickness. If this measurement is not available, output “N/A”.

Field 17, Title “Intraparenchymal hemorrhage volume calculation”: If field 14 is “YES Dimensions listed” and the dimensions include intraparenchymal hemorrhage volume or dimensions that can be used to estimate the volume using the ellipsoid formula, output the numeric value for the volume in mL. If volume is provided with different units then convert to mL. If there are dimensions provided for multiple regions of intraparenchymal hemorrhage, provide the sum of the volumes; if calculating the volume using the ellipsoid formula then the volumes of each region should be calculated separately before adding them together. If these measurements are not available, output “N/A”.

Field 18, Title “Intraparenchymal hemorrhage volume category”. If field 17 contains a numeric value, output the relevant category from the following options: “Small (>0 to <5mL)”, “Medium (≥5 to <20mL)”, “Large (≥20 to <50mL)” and “Very large (≥50mL)”. If field 17 is “N/A”, output “N/A”.

Field 19, Title, “Recent intracranial surgery?”: Output “YES Recent surgery” if the report suggests recent intracranial surgery and, else, output “NO Recent surgery”.

Field 20, Title, “Recent intracranial surgery qualitative”: If field 19 is “YES Recent surgery”, output the phrasing from the report that suggests there was recent intracranial surgery. If field 19 is “NO Recent surgery”, output “N/A”.

Field 21, Title, “Intracranial artifact present?”: Output “YES Intracranial artifact” if there is any intracranial artifact described in the report and, else, output “NO Intracranial artifact”.

Field 22, Title, “Intracranial artifact present qualitative”: If field 21 is “YES Intracranial artifact”, output the specific intracranial artifact described in the report. If there are multiple intracranial artifacts, list the description for each artifact in the same JSON string value as a semicolon-separated list. If field 21 is “NO Intracranial artifact”, output “N/A”.

Field 23, Title, “Intracranial calcification present?”: Output “YES Intracranial calcification” if there is any intracranial calcification described in the report and, else, output “NO Intracranial calcification”.

Field 24, Title, “Intracranial calcification qualitative”: If field 23 is “YES Intracranial calcification”, output the specific intracranial calcification or calcifications described in the report. If there are multiple intracranial calcifications, list the description for each intracranial calcification in the same JSON string value as a semicolon-separated list. If field 23 is “NO Intracranial calcification”, output “N/A”.

Field 25, Title, “Intracranial hyperdense (non-hemorrhagic) lesion present?”: Output “YES Hyperdense (non-hemorrhagic) lesion” if there is an intracranial hyperdense (non-hemorrhagic) lesion described in the report and, else, output “NO Hyperdense (non-hemorrhagic) lesion”.

Field 26, Title, “Intracranial hyperdense (non-hemorrhagic) lesion qualitative”: If field 25 is “YES Hyperdense (non-hemorrhagic) lesion”, output the specific intracranial hyperdense lesion or lesions described in the report. If there are multiple hyperdense lesions, list the description for each hyperdense lesion in the same JSON string value as a semicolon-separated list. If field 25 is “NO Hyperdense (non-hemorrhagic) lesion”, output “N/A”.

Field 27, Title, “Extra-axial (non-hemorrhagic) collection present?”: Output “YES Extra-axial (non-hemorrhagic) collection” if there is an extra-axial (non-hemorrhagic) collection described in the report and, else, output “NO Extra-axial (non-hemorrhagic) collection”.

Field 28, Title, “Extra-axial (non-hemorrhagic) collection qualitative”: If field 27 is “YES Extra-axial (non-hemorrhagic) collection”, output the specific extra-axial collection or collections described in the report. If there are multiple extra-axial collections, list the description for each extra-axial collection in the same JSON string value as a semicolon-separated list. If field 27 is “NO Extra-axial (non-hemorrhagic) collection”, output “N/A”.
